# Temporal Changes in Clinical Practice with COVID-19 Hospitalized Patients: Potential Explanations for Better In-Hospital Outcomes

**DOI:** 10.1101/2020.09.29.20203802

**Authors:** Kevin E. Kip, Graham Snyder, Donald M. Yealy, John W. Mellors, Tami Minnier, Michael P. Donahoe, Jeffrey McKibben, Kevin Collins, Oscar C. Marroquin

**Author notes:** **Corresponding Author:** Kevin E. Kip, Ph.D., FAAAS, FAHA, Vice President of Clinical Analytics | UPMC Health Services Division, 3600 Forbes & Meyran | Forbes Tower, 9th Floor, Suite 10070, Pittsburgh, PA 15213.

## Abstract

**Background/Aims:** We reviewed demographic and clinical profiles, along with measures of hospital-based clinical practice to identify temporal changes in clinical practice that may have affected in-hospital outcomes of patients with COVID-19.

**Methods:** Data consisted of sociodemographic and clinical data captured in University of Pittsburgh Medical Center (UPMC) electronic medical record (EMR) systems, linked by common variables (deidentified). The analysis population included hospitalized patients (across 21 hospitals) with a primary diagnosis of COVID-19 infection during the period March 14-August 31, 2020. The primary outcome was a composite of in-hospital mechanical ventilation/mortality. We compared temporal trends in patient characteristics, clinical practice, and hospital outcomes using 4 time-defined epochs for calendar year 2020: March 14-March 31 (epoch 1); April 1-May 15, (epoch 2), May 16-June 28 (epoch 3); and June 29-August 31 (epoch 4). We report unadjusted survival estimates, followed by propensity score analyses to adjust for differences in patient characteristics, to compare in-hospital outcomes of epoch 4 patients (recently treated) to epoch 1-3 patients (earlier treated).

**Results:** Mean number of hospital admissions was 9.9 per day during epoch 4, which was ∼2-to 3-fold higher than the earlier epochs. Presenting characteristics of the 1,076 COVID-19 hospitalized patients were similar across the 4 epochs, including mean age. The crude rate of mechanical ventilation/mortality was lower in epoch 4 patients (17%) than in epoch 1-3 patients (23% to 35%). When censoring for incomplete patient follow-up, the rate of mechanical ventilation/mortality was lower in epoch 4 patients (*p*<0.0001), as was the individual component of mechanical ventilation (*p*=0.0002) and mortality (*p*=0.02). In propensity score adjusted analyses, the in-hospital relative risk (RR) of mechanical ventilation/mortality was lower in epoch 4 patients (RR=0.67, 95% CI: 0.48, 0.93). For the outcome being discharged alive within 3, 5, or 7 days of admission, adjusted odds ranged from 1.6-to 1.7-fold higher among epoch 4 patients compared to earlier treated patients. The better outcomes in epoch 4 patients were principally observed in patients under the age of 75 years. Patient level dexamethasone use was 55.6% in epoch 4 compared to 15% or less of patients in the earlier epochs. Most patients across epochs received anticoagulation drugs (principally heparin). Overall steroid (81.7% vs. 54.3%, *p*<0.0001) and anticoagulation use (90.4% vs. 80.7%, *p*=0.0001) was more frequent on the day or day after hospitalization in epoch 4 patients compared to earlier treated patients.

**Conclusions:** In our large system, recently treated hospitalized COVID-19 patients had lower rates of in-hospital mechanical ventilation/mortality and shorter length of hospital stay. Alongside of this was a change to early initiation of glucocorticoid therapy and anticoagulation. The extent to which the improvement in patient outcomes was related to changes in clinical practice remains to be established.

## INTRODUCTION

Since the onset of the COVID-19 pandemic, the US and countries worldwide have seen variation in reported incidence, testing patterns, case fatality rates, investigations of novel treatments, and clinical practice approaches. A few,^1-3^ but not all,^4^ recent non-peer reviewed reports suggested that the case fatality rate of COVID-19 infection is decreasing, and care is changing. However, there is an absence of published reports on large series of hospitalized COVID-19 patients, particularly with respect to temporal changes in clinical outcomes within the same data system.

One key intervention is mechanical ventilation, initially thought to be best started early with more severe COVID-19 respiratory finding, notably hypoxemia. Over time, informal reports note less mechanical ventilation use; potential explanations for lower rates of COVID-19 mechanical ventilation and less mortality include: (i) changing demographics of patients;^5^ (ii) more judicious use of starting and using mechanical ventilation;^6^ (iii) more frequent use of anticoagulants;^7,8^ and steroids;^9-12^ (iv) changing prominent disease manifestations of patients;^13^ (v) seasonal effects of temperature and humidity variation;^14^ and (vi) potential changes in viral infectivity or pathogenicity.^15,16^

There have been temporal changes in COVID-19 testing in addition to no national testing strategy. Early in the pandemic, only those with symptoms and a higher pretest probability of COVID-19 exposure or disease were the focus of testing. Soon thereafter, COVID-19 testing increased in many, notably populations with higher frequency of potential infection (e.g., nursing home patients as well as health care professionals), and more recently increased screening of asymptomatic patients such as those with return to college campus activities, along with more intensive contacting tracing (and testing) of individuals likely to have been exposed to those carrying SARS CoV2 virus.^17-20^

We sought to examine clinical outcomes of hospitalized COVID-19 patients in a large health care system accompanied by an assessment of demographic and clinical profiles, along with changes in hospital-based clinical practice, that have occurred since the onset of the COVID-19 pandemic in the US.

## METHODS

### Sources of Data

We used data routinely captured in the University of Pittsburgh Medical Center (UPMC) electronic medical record (EMR) systems. In brief, UPMC is a large academic medical center and insurer, housed principally in Pennsylvania.^21^ The UPMC data system has detailed sociodemographic and medical history data, diagnostic and clinical tests conducted, surgical and other treatment procedures performed, prescriptions ordered, and billing charges on all outpatient and in-hospital encounters, with diagnoses and procedures coded based on the International Classification of Diseases, Ninth and Tenth revisions (ICD-9 and ICD-10, respectively).

We linked the primary data sources using common variables (deidentified) within the UPMC data ecosystem aggregated in its Clinical Data Warehouse (CDW) that include: (i) *Medipac*, the admit, discharge and transfer registration and hospital-based billing system; (ii) *Cerner*, the inpatient electronic medical record (EMR) for relevant clinical information for bedded patients at a UPMC inpatient hospital; (iii) *Epic*, the UPMC EMR for ambulatory office visits owned by UPMC (Community Medicine Inc. and (University of Pittsburgh Physicians); and (iv) *Aria*, the EMR utilized in most ambulatory Cancer Centers at UPMC for both radiation oncology and medical oncology. In calendar year 2019 among discharged UPMC hospital patients, there were 306,456 visits among 201,829 unique patients, with mean (median) age of 54 (60) years, 54% female, and median length of stay of 2.6 days.

### Patient Population

We studied 139,465 patients with nucleic acid amplification tests for SARS-CoV-2 (the cause of COVID-19) during the period March 14, 2020 to August 31, 2020. Of these, the study population consisted of 1,076 patients who tested positive for COVID-19 and were hospitalized at one of 21 UPMC hospitals (see **Supplemental Table S1** for listing of hospitals). Our study received formal ethics approval by the UPMC Ethics and Quality Improvement Review Committee (Project ID 2882), the ethics/oversight body for ensuring patient confidentiality and consent (including waiver of consent) for analysis and dissemination of deidentified data within the UPMC system.

### Primary and Secondary Outcomes

The primary outcome for this analysis was a composite of in-hospital mechanical ventilation or mortality. We assessed in-hospital mechanical by the presence of a charge of mechanical ventilation within the Medipac billing software, specifically codes 94002 (first day of mechanical ventilation) and 94003 (each subsequent day of mechanical ventilation). Because some facilities used intensive care units for the care of COVID-19 patients regardless of illness acuity, we did not use intensive care unit admission as an outcome. We assessed in-hospital mortality by the discharge disposition of “Ceased to Breathe” sourced from the inpatient Medical Record System.

Secondary outcomes included in-hospital mechanical ventilation or mortality individually and length of hospital stay (calculated by taking the difference between a discharge date time and the admission date time) available through billing within the Medipac system. Study investigators remained unaware of ascertainment of mechanical ventilation and mortality within the UPMC system during data collection and analysis.

### Explanatory Variables

For assessment of temporal changes and prior to analyses, we categorized the study analysis period into discrete epochs based on observed empirical changes in testing patterns in the UPMC system. We chose a 4-time period classification scheme (see **Figure 1**), (*Epoch 1*: March 14-March 31, 2020; *Epoch 2*: April 1-May 15, 2020; *Epoch 3*: May 16-June 28, 2020; *Epoch 4*: June 29-August 31, 2020), and then collapsed the 4 epochs into 2 epochs to depict earlier (*Epochs 1,2,3*) versus recently admitted hospital patients (*Epoch 4*). For assessment of variation between epochs, we considered demographic variables, clinical history and medical comorbidities, laboratory values, vital signs, and medication use, with a focus on indicators of changing clinical practice such as use and timing of specific medications. As above, investigators were unaware of initial documentation of the explanatory variables in the EMRs.

**Figure 1.**
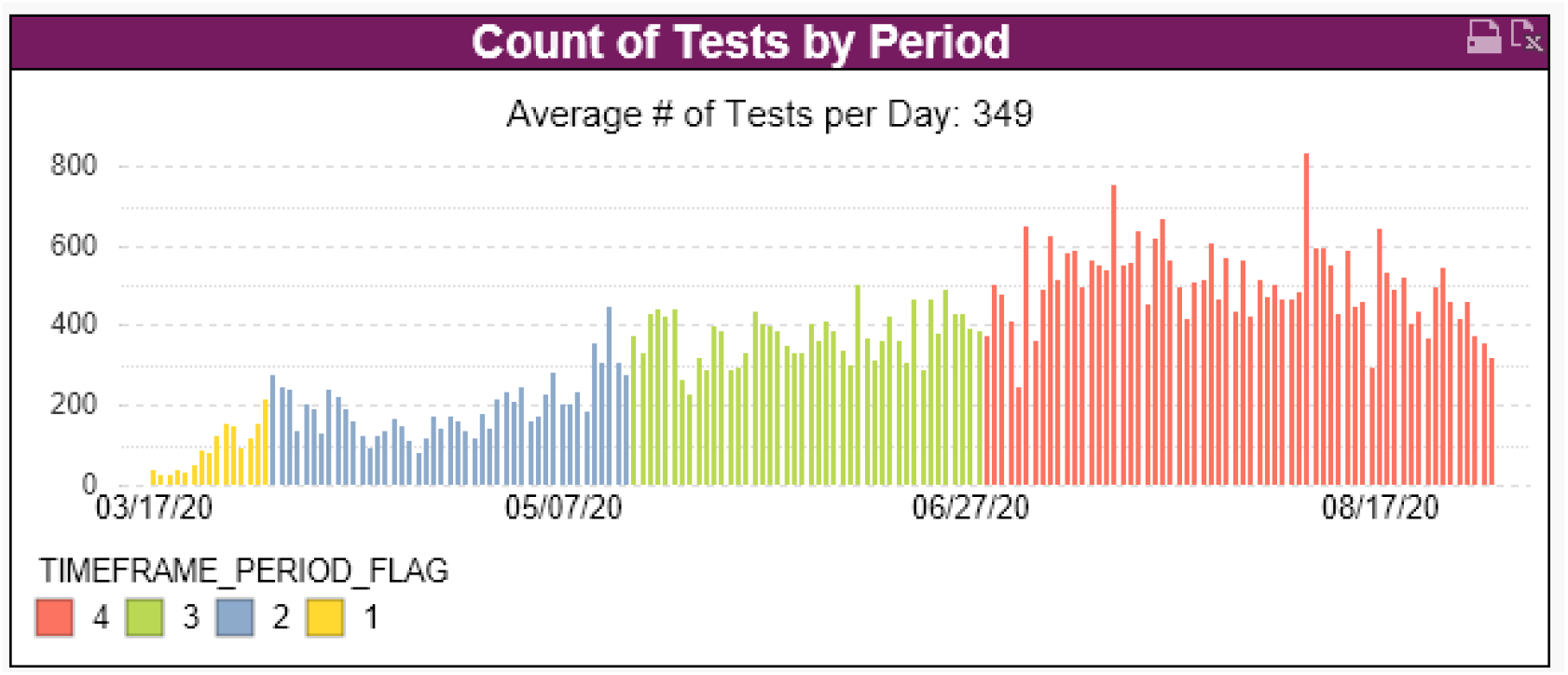
Histogram of the average number of COVID-19 tests reported in the UPMC system during the study period. The color coding depicts the time periods in which the 4 epochs were defined for comparative analyses.

### Statistical Methods

We compared characteristics of patients across the 4 epochs using analysis of variance (ANOVA) for continuous variables and chi-square tests for categorical variables. We used the same methods to compare patterns of use of drugs and unadjusted in-hospital outcomes by epoch group, while using non-parametric Wilcoxon tests used for variables with skewed distributions, in particular, days from hospital admission to initiation of specific drugs or oxygen therapies. We defined 3 binary “shorter” hospital length of stay variables for patients discharged alive as ≤3, ≤5, and ≤7 days, and included only patients admitted with adequate time to meet these definitions. We used chi-square tests to compare proportions by epochs. Then, we used Kaplan-Meier survival curves to present the outcomes in-hospital mechanical ventilation/mortality and each individual component, comparing epoch 4 (recent) versus epoch 1-3 (earlier) patients by use of the log-rank test.

We used propensity score methodology to compare in-hospital outcomes between epoch 4 versus epoch 1-3, adjusting for differences in presenting characteristics,.^22,23^ Logistic regression models were fit using hospital admission during epoch 4 as the dependent variable with stepwise selection (at *p* < 0.2) of measured pre-treatment explanatory variables. Resulting propensity scores (i.e. predicted probability of being in epoch 4 versus epochs 1-3) were the output and used as a continuous variable to control for confounding in Cox proportional hazards regression and logistic regression models of in-hospital outcomes. In sensitivity analyses, models were replicated with the use of inverse probability weighting (IPW), as well as 1:1 propensity score matching (PSM) with a maximum propensity score probability difference of 0.01. We set the comparison alpha error at 0.05 without correction for multiple comparisons.

In stratified analyses, we examined potential effect modification by patient age using 3 groups defined as: (i) less than 60 years of age; (ii) 60 to less than 75 years of age; and (iii) 75 years of age and older. We did not impute missing values in any of the analyses. Methods and results are reported in accordance with The REporting of studies Conducted using Observational Routinely-collected health Data (RECORD) statement^24^ (see **Supplemental Table 2)**.

## RESULTS

The mean number of hospital admissions was 9.9 per day during epoch 4, which was 2-to 3-fold higher than average daily admission during the earlier epochs (see **Supplementary Figure S1**). The overall rate of COVID-19 testing performed in epoch 4 also was higher (**Figure 1**).

Presenting characteristics of patients hospitalized with COVID-19 are listed in **Table 1**. The mean patient age was similar across the 4 epochs (ranging from 63.0 to 65.2 years), as was representation by sex and race. In the earliest epoch (1) and most current epoch (4), the proportion of patients covered by UPMC health insurance was higher. Otherwise, the majority of patient characteristics were similar across the four epochs. Exceptions included a higher prevalence of COPD in epoch 2 and 4 patients, and nominally different mean hemoglobin laboratory values. Recent outpatient medication use at the time of hospital admission was similar across the 4 epochs.

**Table 1.**
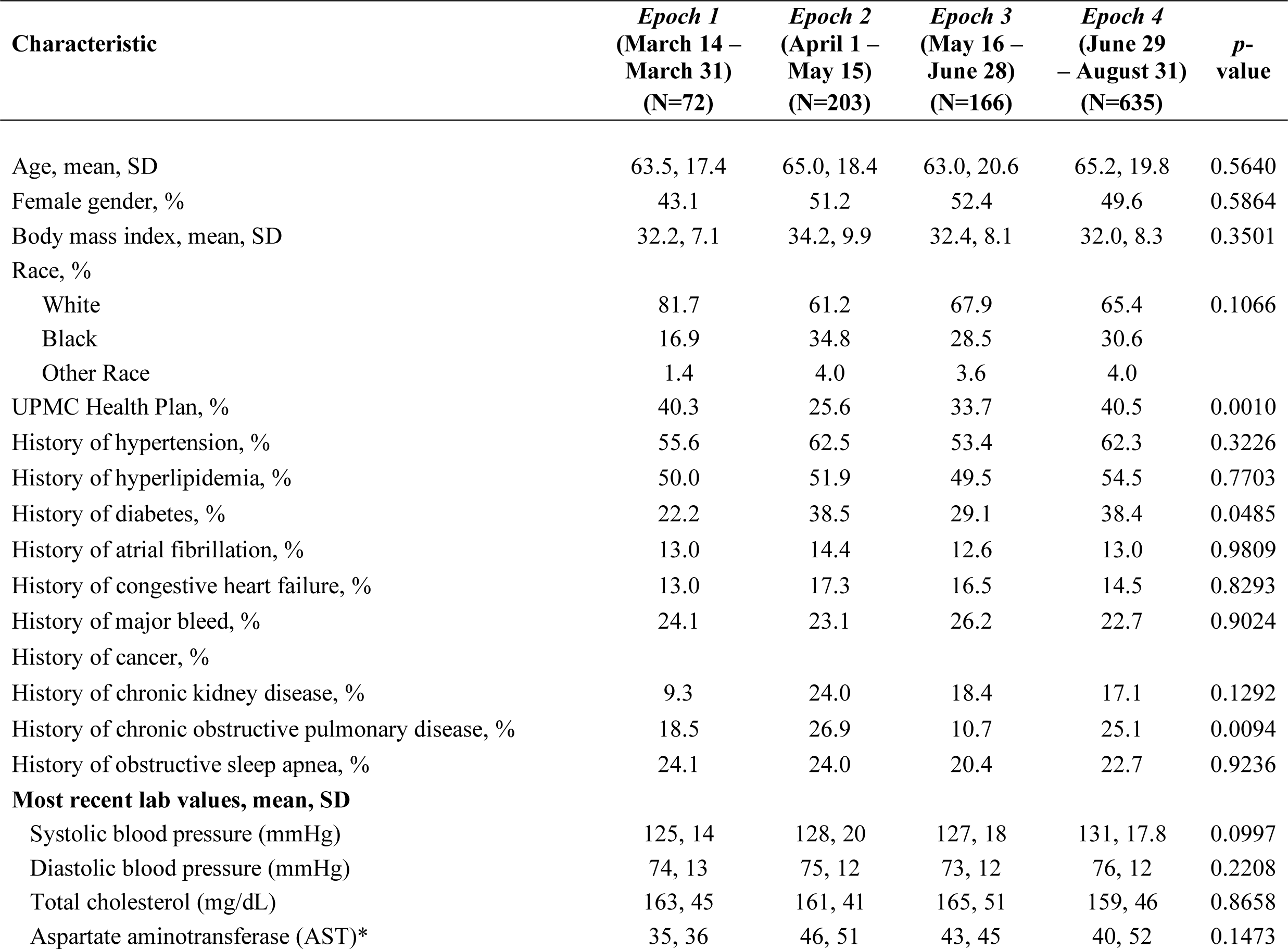
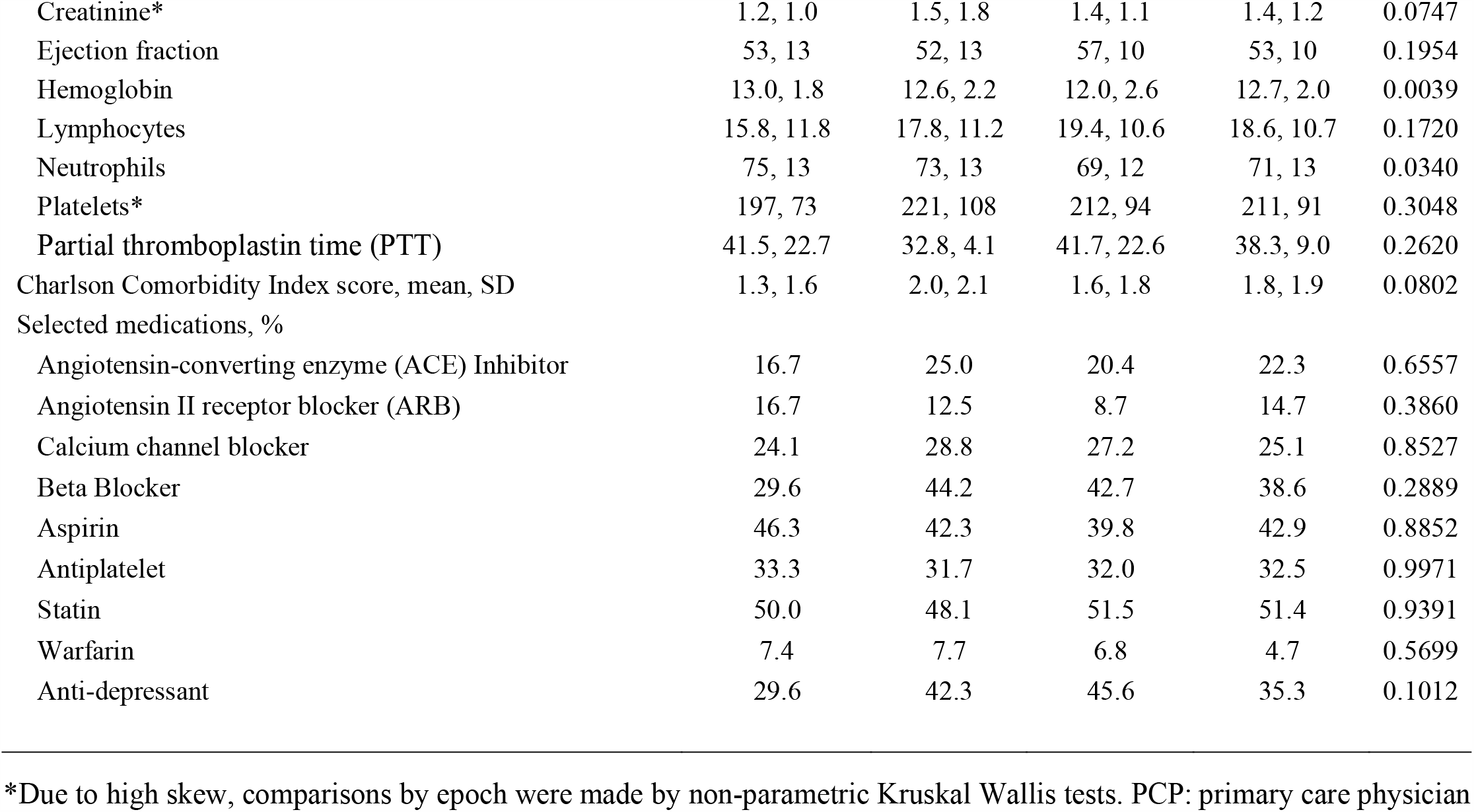
Comparison of Hospitalized COVID Patient Characteristics at Admission by Epoch

| **Characteristic** | ***Epoch 1***<br>**(March 14 –**<br>**March 31)**<br>**(N=72)** | ***Epoch 2***<br>**(April 1 –**<br>**May 15)**<br>**(N=203)** | ***Epoch 3***<br>**(May 16 –**<br>**June 28)**<br>**(N=166)** | ***Epoch 4***<br>**(June 29**<br>**– August 31)**<br>**(N=635)** | ***p*-**<br>**value** |
| --- | --- | --- | --- | --- | --- |
| Age, mean, SD | 63.5, 17.4 | 65.0, 18.4 | 63.0, 20.6 | 65.2, 19.8 | 0.5640 |
| Female gender, % | 43.1 | 51.2 | 52.4 | 49.6 | 0.5864 |
| Body mass index, mean, SD | 32.2, 7.1 | 34.2, 9.9 | 32.4, 8.1 | 32.0, 8.3 | 0.3501 |
| Race, % |  |  |  |  |  |
| White | 81.7 | 61.2 | 67.9 | 65.4 | 0.1066 |
| Black | 16.9 | 34.8 | 28.5 | 30.6 |  |
| Other Race | 1.4 | 4.0 | 3.6 | 4.0 |  |
| UPMC Health Plan, % | 40.3 | 25.6 | 33.7 | 40.5 | 0.0010 |
| History of hypertension, % | 55.6 | 62.5 | 53.4 | 62.3 | 0.3226 |
| History of hyperlipidemia, % | 50.0 | 51.9 | 49.5 | 54.5 | 0.7703 |
| History of diabetes, % | 22.2 | 38.5 | 29.1 | 38.4 | 0.0485 |
| History of atrial fibrillation, % | 13.0 | 14.4 | 12.6 | 13.0 | 0.9809 |
| History of congestive heart failure, % | 13.0 | 17.3 | 16.5 | 14.5 | 0.8293 |
| History of major bleed, % | 24.1 | 23.1 | 26.2 | 22.7 | 0.9024 |
| History of cancer, % |  |  |  |  |  |
| History of chronic kidney disease, % | 9.3 | 24.0 | 18.4 | 17.1 | 0.1292 |
| History of chronic obstructive pulmonary disease, % | 18.5 | 26.9 | 10.7 | 25.1 | 0.0094 |
| History of obstructive sleep apnea, % | 24.1 | 24.0 | 20.4 | 22.7 | 0.9236 |
| **Most recent lab values, mean, SD** |  |  |  |  |  |
| Systolic blood pressure (mmHg) | 125, 14 | 128, 20 | 127, 18 | 131, 17.8 | 0.0997 |
| Diastolic blood pressure (mmHg) | 74, 13 | 75, 12 | 73, 12 | 76, 12 | 0.2208 |
| Total cholesterol (mg/dL) | 163, 45 | 161, 41 | 165, 51 | 159, 46 | 0.8658 |
| Aspartate aminotransferase (AST)* | 35, 36 | 46, 51 | 43, 45 | 40, 52 | 0.1473 |
| Creatinine* | 1.2, 1.0 | 1.5, 1.8 | 1.4, 1.1 | 1.4, 1.2 | 0.0747 |
| Ejection fraction | 53, 13 | 52, 13 | 57, 10 | 53, 10 | 0.1954 |
| Hemoglobin | 13.0, 1.8 | 12.6, 2.2 | 12.0, 2.6 | 12.7, 2.0 | 0.0039 |
| Lymphocytes | 15.8, 11.8 | 17.8, 11.2 | 19.4, 10.6 | 18.6, 10.7 | 0.1720 |
| Neutrophils | 75, 13 | 73, 13 | 69, 12 | 71, 13 | 0.0340 |
| Platelets* | 197, 73 | 221, 108 | 212, 94 | 211, 91 | 0.3048 |
| Partial thromboplastin time (PTT) | 41.5, 22.7 | 32.8, 4.1 | 41.7, 22.6 | 38.3, 9.0 | 0.2620 |
| Charlson Comorbidity Index score, mean, SD | 1.3, 1.6 | 2.0, 2.1 | 1.6, 1.8 | 1.8, 1.9 | 0.0802 |
| Selected medications, % |  |  |  |  |  |
| Angiotensin-converting enzyme (ACE) Inhibitor | 16.7 | 25.0 | 20.4 | 22.3 | 0.6557 |
| Angiotensin II receptor blocker (ARB) | 16.7 | 12.5 | 8.7 | 14.7 | 0.3860 |
| Calcium channel blocker | 24.1 | 28.8 | 27.2 | 25.1 | 0.8527 |
| Beta Blocker | 29.6 | 44.2 | 42.7 | 38.6 | 0.2889 |
| Aspirin | 46.3 | 42.3 | 39.8 | 42.9 | 0.8852 |
| Antiplatelet | 33.3 | 31.7 | 32.0 | 32.5 | 0.9971 |
| Statin | 50.0 | 48.1 | 51.5 | 51.4 | 0.9391 |
| Warfarin | 7.4 | 7.7 | 6.8 | 4.7 | 0.5699 |
| Anti-depressant | 29.6 | 42.3 | 45.6 | 35.3 | 0.1012 |

\*Due to high skew, comparisons by epoch were made by non-parametric Kruskal Wallis tests. PCP: primary care physician

**Table 2.** Practice Patterns of Use of Drugs by Epoch*

| Characteristic | March 14 –<br>March 31<br>(N=72) | April 1 –<br>May 15<br>(N=203) | May 16 –<br>June 28<br>(N=166) | June 29 –<br>August 31<br>(N=635) | p-value |
| --- | --- | --- | --- | --- | --- |
| Use of remdesivir, % | 0.0 | 3.9 | 31.3 | 17.8 | <.0001 |
| Use of dexamethasone, % | 5.6 | 5.4 | 14.5 | 55.6 | <.0001 |
| Use of hydrocortisone, % | 2.8 | 11.3 | 6.6 | 3.3 | <.0001 |
| Use of prednisone, % | 8.3 | 6.4 | 7.2 | 6.8 | 0.9499 |
| Use of methylprednisolone, % | 8.3 | 10.3 | 9.6 | 9.0 | 0.931 |
| Any steroid used, % | 20.8 | 25.6 | 31.3 | 62.2 | <.0001 |
| Use of heparin, % | 68.1 | 73.4 | 71.1 | 79.2 | 0.0285 |
| Use of anticoagulation, % | 73.6 | 83.3 | 75.3 | 86.5 | 0.0007 |
| Days from admission to remdesivir, mean, SD | ., . | 4.8, 2.9 | 2.2, 3.8 | 1.7, 2.4 | 0.3734 |
| Days from admission to dexamethasone, mean, SD | 3.8, 6.2 | 7.0, 8.0 | 4.6, 7.6 | 1.5, 3.1 | 0.0135 |
| Days from admission to hydrocortisone, mean, SD | 14.5, 10.6 | 5.7, 6.7 | 1.6, 1.4 | 7.5, 7.8 | 0.6182 |
| Days from admission to prednisone, mean, SD | 14.2, 21.8 | 2.9, 4.6 | 3.8, 3.3 | 4.3, 7.1 | 0.4186 |
| Days from admission to methylprednisolone, mn, SD | 8.5, 18.4 | 5.9, 7.4 | 3.3, 4.8 | 4.4, 8.9 | 0.1329 |
| Days from admission to steroid use, mean, SD | 5.0, 7.9 | 4.3, 6.5 | 2.6, 3.8 | 1.3, 2.9 | <.0001 |
| Days from admission to heparin, mean, SD | 1.8, 3.0 | 1.5, 5.7 | 1.2, 2.4 | 1.0, 3.2 | 0.0003 |
| Days from admission to anticoagulation, mean, SD | 1.6, 2.9 | 1.0, 1.3 | 1.2, 2.3 | 0.8, 1.6 | 0.0005 |

\*For all days from admission variables, comparisons by epoch were made by non-parametric Wilcoxon continuity-corrected 2-sample tests comparing epochs 1-3 to epoch 4.

### Prescribing Patterns for Inpatients

Reported timing and use of medications showed changing patterns across the 4 epochs (**Table 2**). Specifically, the use of remdesivir peaked in epoch 3 at 31.3% and then declined to 17.8% in epoch 4. There was an increase in the use of dexamethasone to 55.6% of patients in epoch 4 compared to 15% or less of patients in the earlier epochs. The majority of patients received anticoagulation drugs, principally heparin, and use was highest in epoch 2 and 4 patients. There was a striking difference in the timing of drug initiation, with much earlier use of dexamethasone, glucocorticoids as a class, and anticoagulation in epoch 4 patients. As depicted in **Figure 2**, a higher percentage of patients in epoch 4 had initiation of steroids (*p*<0.0001) or anticoagulation (*p*=0.0001) on the same day (Day 0) or following day (Day 1) after hospital admission compared to patients admitted in epochs 1-3.

**Figure 2.**
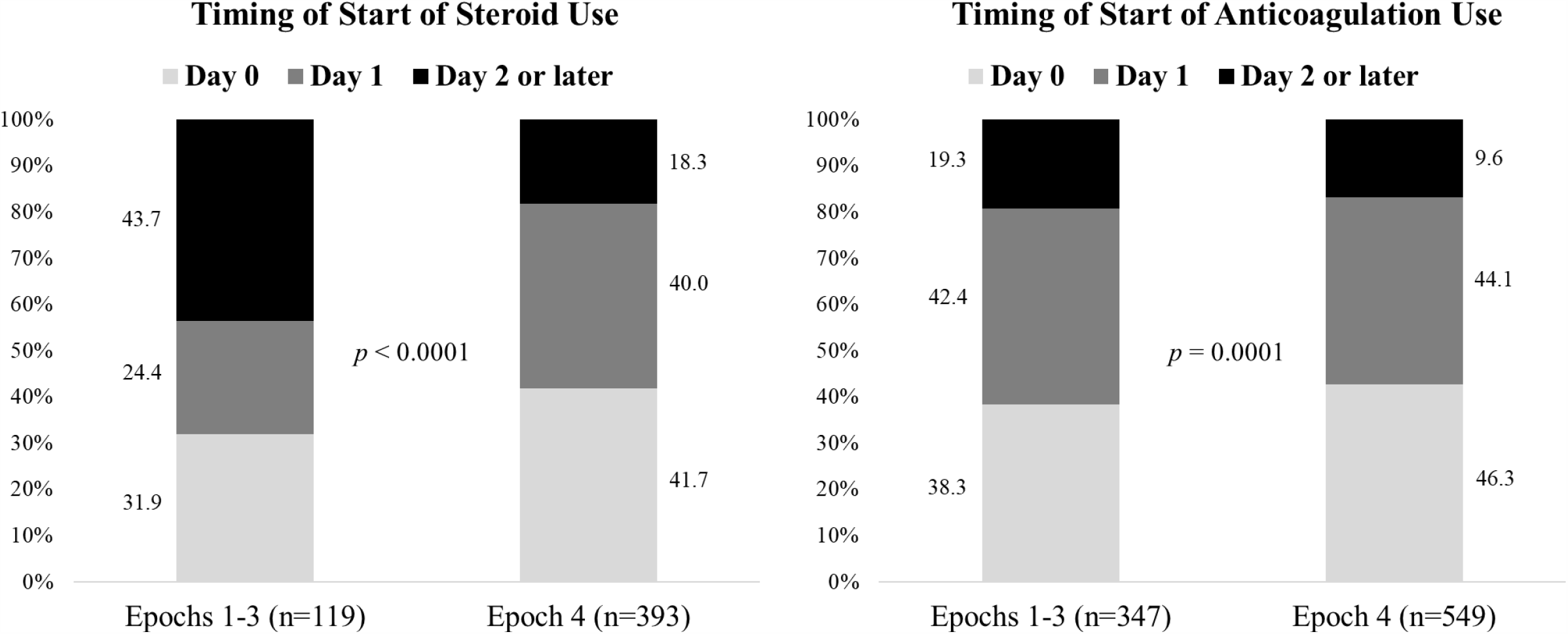
Stacked bar chart of timing of use and steroids and anticoagulants comparing recently treated epoch 4 patients to earlier patients treated in epochs 1-3. Light shading: drug initiated on same day as hospital admission; Medium shading: drug administered on the day after hospital admission; Dark shading: drug administered 2 or more days after hospital admission.

### Use of Oxygen Support and In-Hospital Outcomes

More than half of all patients received oxygen therapy during their hospital stay, with the highest percentage (73%) observed in epoch 2 patients (**Table 3**). Oxygen therapy started sooner in epoch 4 patients (mean of 1.9 days), and initiation of drug aerosol therapy was also sooner in epoch 4 patients (mean of 4.0 days) compared to epoch 1-3 patients (mean of 5.6 to 9.2 days). By contrast, the timing of the start of mechanical ventilation after hospital admission was longest in epoch 4 patients (mean of 3.6 days) compared to epoch 1-3 patients (mean of 1.9 to 2.3 days). Individually, the crude rate of use of mechanical ventilation was lower in epoch 4 patients (11.7%) than in epoch 1-3 patients (17% to 22%) as was in-hospital mortality (10.2% vs. 14.5% to 23.6%), recognizing that not all epoch 4 patients had been discharged. For the primary outcome, the crude rate of mechanical ventilation/mortality was lower in epoch 4 patients (17%) than in epoch 1-3 patients (23% to 35%). When censoring for incomplete patient follow-up (i.e. patients admitted yet not yet discharged), rates of mechanical ventilation (*p*=0.0002), mortality (*p*=0.02), and mechanical ventilation/mortality (*p*<0.0001) were lower in epoch 4 patients compared to epoch 1-3 patients (**Supplemental Figures 2a, 2b, 2c**, respectively). Rates of being discharged alive within 3, 5, and 7 days of admission were significantly higher in epoch 4 patients compared to epoch 1-3 patients (**Table 3**).

**Table 3.**
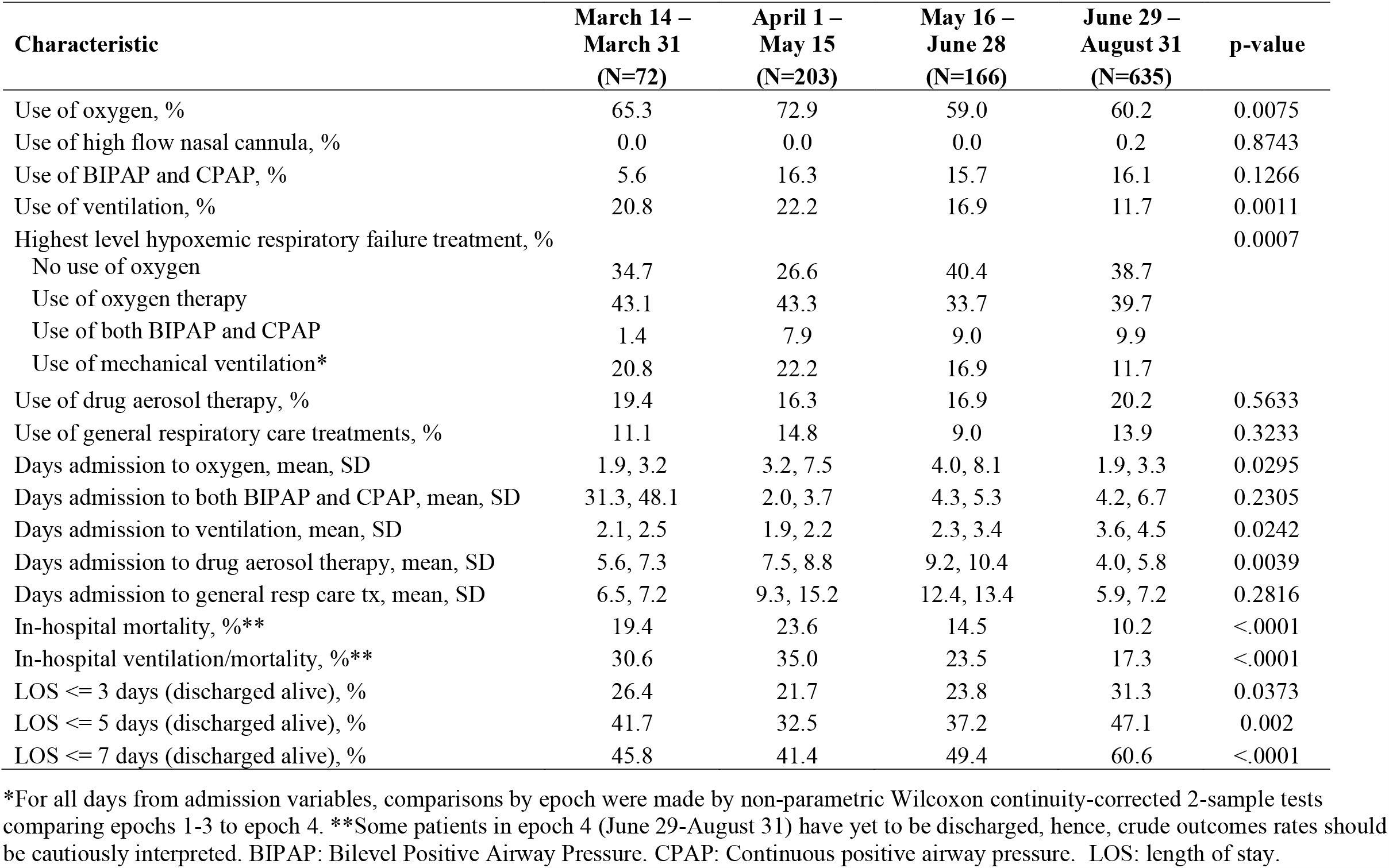
Practice Patterns of Use of Oxygen Support and Hospital Outcomes by COVID Epoch*

### Relative Risks of In-Hospital Outcomes

In propensity score adjusted Cox regression models, the estimated in-hospital relative risks (RR: expressed as hazard ratios) of the composite outcome mechanical ventilation/mortality (RR=0.67, 95% CI: 0.48, 0.93), use of mechanical ventilation (RR=0.64, 95% CI: 0.42, 0.96), and mortality, (RR=0.81, 95% CI: 0.53, 1.23) were lower in epoch 4 patients compared to epoch 1-3 patients (**Table 4**). In propensity adjusted logistic regression models, RRs (expressed as odds ratios) of being discharged alive within 3, 5, or 7 days of admission were higher in epoch 4 patients compared to epoch 1-3 patients, ranging from an estimated 1.6-to 1.7-fold higher odds of shorter length of stay in epoch 4 patients (**Table 4**).

**Table 4.**
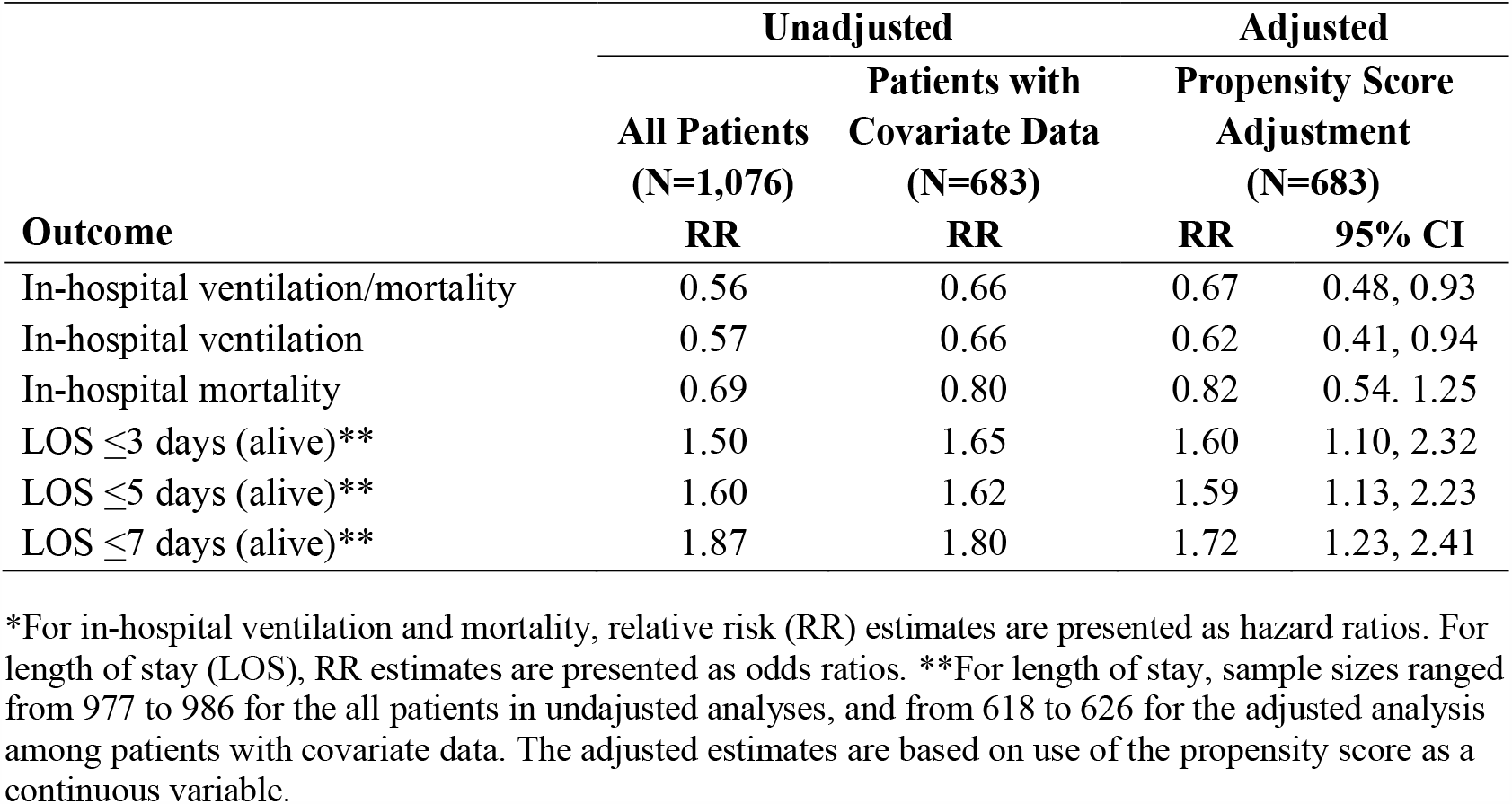
Relative Risks* of In-Hospital Outcomes Comparing Epoch 4 (Recent) to Epoch 1/2/3 (Earlier) Treated COVID-19 Patients

In sensitivity analyses using 1:1 propensity matched group patients (epoch 4 vs. epochs 1-3), presenting patient characteristics were similar (see **Supplemental Table S3**), and Cox and logistic regression estimates favoring better in-hospital outcomes in epoch 4 patients were similar (see **Supplementary Table S4**). Outcome results using propensity inverse probability weighting **Supplementary Table S4**) were similar. A comparison of outcome results across all methods of analysis is depicted in **Supplemental Figure 2**, which shows similar results across all methods.

### Relative Risks of In-Hospital Outcomes by Age

As seen in **Table 5**, the in-hospital outcome changes observed in epoch 4 versus epoch 1-3 patients varied by age. Specifically, in the age groups of less than 60 years of age or 60 to 74 years of age, the adjusted risk of in-hospital mechanical ventilation/mortality was approximately 50% lower in epoch 4 patients compared to epoch 1-3 patients. In contrast, the adjusted risk of in-hospital mechanical ventilation/mortality among patients 75 years of age and older was similar between epoch 4 versus epoch 1-3 patients.

**Table 5.** Relative Risks* of In-Hopsital Outcomes Comparing Epoch 4 (Current) to Epoch 1/2/3 (Earlier) COVID-19 Patients by Age Groups

| Outcome / Age Group | N | # Events | Propensity Score Adjustment |  |
| --- | --- | --- | --- | --- |
|  |  |  | RR | 95% CI |
| Mechanical ventilation/mortality |  |  |  |  |
| Less than 60 years of age | 233 | 30 | 0.45 | 0.22, 0.95 |
| 60 to less than 75 years of age | 207 | 54 | 0.52 | 0.29, 1.93 |
| 75 years of age and older | 243 | 71 | 0.99 | 0.58, 1.66 |
| Mechanical ventilation |  |  |  |  |
| Less than 60 years of age | 233 | 28 | 0.44 | 0.21, 0.95 |
| 60 to less than 75 years of age | 207 | 40 | 0.55 | 0.28, 1.07 |
| 75 years of age and older | 243 | 34 | 1.14 | 0.51, 2.55 |
| In-hospital mortality |  |  |  |  |
| Less than 60 years of age | 233 | 9 | 0.59 | 0.14, 2.41 |
| 60 to less than 75 years of age | 207 | 32 | 0.47 | 0.21, 1.05 |
| 75 years of age and older | 243 | 58 | 0.91 | 0.52, 1.62 |
| LOS $\leq 3$ days (alive) | | | | |
| Less than 60 years of age | 217 | 104 | 1.81 | 1.04, 3.15 |
| 60 to less than 75 years of age | 188 | 43 | 1.80 | 0.85, 3.83 |
| 75 years of age and older | 221 | 43 | 1.23 | 0.56, 2.67 |
| LOS $\leq 5$ days (alive) | | | | |
| Less than 60 years of age | 215 | 137 | 2.14 | 1.20, 3.79 |
| 60 to less than 75 years of age | 188 | 67 | 1.23 | 0.64, 2.35 |
| 75 years of age and older | 220 | 80 | 1.45 | 0.77, 2.73 |
| LOS $\leq 7$ days (alive) | | | | |
| Less than 60 years of age | 214 | 154 | 2.63 | 1.42, 4.88 |
| 60 to less than 75 years of age | 186 | 94 | 1.90 | 1.01, 3.55 |
| 75 years of age and older | 218 | 95 | 1.23 | 0.67, 2.26 |

\*For in-hospital ventilation and mortality, relative risk (RR) estimates are presented as hazard ratios. For length of stay (LOS), RR estimates are presented as odds ratios. The adjusted estimates are based on use of the propensity score as a continuous variable.

Among patients less than 60 years of age, the adjusted odds of having a hospital stay of ≤3 days (discharged alive) were approximately 1.8 fold higher in epoch 4 patients, and were even larger for a hospital stay of ≤ 5 days (about 2.1-fold higher) or ≤ 7 days (about 2.6-fold higher). Patients age 60 to less than 75 years also tended to have a shorter length of stay in epoch 4, whereas there were no differences by epoch in lengh of for patients aged 75 and older.

## DISCUSSION

Among COVID-19 patients hospitalized starting in March of 2020, our analyses show recent (epoch 4: June 19^th^ to August 31^st^, 2020) decreases in rates of mechanical ventilation, mortality, and hospital length of stay, compared with earlier time periods. These results were observed principally in hospitalized patients under the age of 75, and are consistent with some,^1-3,39-42^ but not all,^4^ recent non-peer-reviewed reports. The better hospital outcomes observed do not appear to be attributable to differences in baseline characteristics of hospitalized COVID-19 patients, which were minimal and controlled for statistically with multiple analytic approaches.

In our large, representative health care system, we observed marked temporal changes in clinical management of patients, The most notable changes in clinical management of patients in the most recent interval was s higher use of dexamethasone (and glucocorticoids overall), higher use of anticoagulants, earlier initiation of dexamethasone, glucocorticoids as a class, and use of anticoagulants. The observed treatment strategy of earlier and significantly more frequent use of dexamethasone in patients in the UPMC system likely was triggered by the July 2020 release of the RECOVERY trial,^9^ which showed lower 28-day mortality in patients who were receiving oxygen with or without invasive mechanical ventilation, and who received dexamethasone as compared to placebo. Our observed practice pattern of more frequent use of steroids is also consistent with a recently published (September 2, 2020) meta-analysis of six trials involving random assignment of different steroids (dexamethasone, hydrocortisone, methylprednisolone) compared to placebo, and approximately 30% lower risk of 28-day mortality among patients treated with steroids.^25^

### Effect of Treatment Changes in Clinical Practice

An obvious question that arises from the present analysis is to what extent did the recent changes in clinical practice (e.g. greater use and earlier initiation of steroids and anticoagulants) lead to overall lower rates of mechanical ventilation and hospital mortality, as well shorter length of hospital stay? Unfortunately, this type of question is difficult to answer with an observational dataset. Specifically, in non-randomized settings, there is potential confounding by indication (indication bias) when comparing different treatment approaches. It is nearly impossible to determine whether a given treatment approach was initiated *a priori*, as opposed to preferentially in response to patient disease severity and/or hospital course, thereby rendering comparisons of such treated patients to those who did not receive the treatment as potentially biased.^26,27^ As recently articulated, we believe caution is wise and causality uncertain though possible with these observational data.^28^ Our data provide a rationale for potential conduct of new RCTs, particularly pragmatic and adaptive types^29^ of trials that can quickly study questions, such as the timing in which steroids and anticoagulants are administered among COVID-19 hospitalized patients.

### Variation in Outcomes by Age

The present analysis indicated that the better in-hospital outcomes observed in recent hospitalized COVID-19 patients (epoch 4) were principally evident in patients under the age of 75, and largely not evident among those 75 years of age and older. Multiple potential explanations exist for these apparent differential results. First may be the clinical decision for potential use of mechanical ventilation. A report of 5,700 COVID-19 patients admitted to 12 hospitals in New York showed high rates of mortality for mechanically ventilated patients over the age of 65.^30^ These findings comport with indications that some physicians may be reluctant to initiate mechanical ventilation in elderly patients as compared to younger patients who typically present with fewer comorbidities, (e.g.^31^) potential medical ethics arguments made for rationing use of ventilators by age,^32,33^ and from the patient perspective, possibility that some elderly patients (coupled with family member input) may be particularly likely to expressly state orders against the use of mechanical ventilation. (e.g.^34-36^) These types of competing influences may blur assessment of the true clinical risk of mechanical ventilation in elderly COVID-19 hospitalized patients. With respect to less indication in our dataset of a mortality reduction in recently treated elderly COVID-19 patients, these findings may represent the greater challenge and complexity in treating elderly hospitalized COVID-19 patients who may be frail and have extensive comorbidities. This is consistent with reports of age being an independent risk factor for COVID-19 mortality^37^ as well as for higher levels of inflammatory dysfunction markers and weakened immune response^38^.

### Strengths and Limitations

Strengths of this study include analysis of a large, heterogenous patient population (e.g. enhances generalizability), standardized collection of real-world data and algorithmic coding of variables harmonized in a clinical data warehouse collected for non-research purposes (e.g. reduces potential reporting and ascertainment bias), and access to a very large battery of sociodemographic, medical history, medication use, and clinical practice and outcome variables available for analysis. A limitation is that we extracted all variables from the EHR of a multisite health care system, making fidelity concerns persist. All current data are observational and cannot determine causality. We also did not collect data to inform changes in host biology or viral pathogenesis over time, and we did not attempt to assess other external factors, such as seasonal effects of temperature and humidity variation, and possible patient-specific directives against the use of mechanical ventilation. Lastly, the present analysis includes a small percentage of hospitalized patients (<13%) enrolled in clinical trials, including potential randomization to either hydroxychloroquine, steroids, immunomodulators, convalescent plasma, or placebo. While a potential impact, we think that effect is modest if at all present.

## Conclusions

Recently treated hospitalized COVID-19 patients in our large health care system have overall lower rates of mechanical ventilation/in-hospital mortality and shorter length of hospital stay compared to earlier intervals. The extent to which these improved patient outcomes are related to recent changing clinical practice (i.e. greater and earlier use of steroids and anticoagulants) is unknown but an opportunity for rigorous future controlled trials.

## Data Availability

The data used in this analysis are not available to outside investigators.

## Figure Legends

**Supplemental Figure 1**.

Bar chart of the average number of hospital admissions during the four different study-defined Epochs: *Epoch 1*: March 14-March 31, 2020; *Epoch 2*: April 1-May 15, 2020; *Epoch 3*: May 16-June 28, 2020; *Epoch 4*: June 29-August 31, 2020.

**Supplemental Figure 2**.

Kaplan-Meier plots of survival free probabilities of in-hospital mechanical ventilation (**2a**), mortality (**2b**), and mechanical ventilation/mortality (**2c**) stratified by epoch 1-3 patients (blue line) versus epoch 4 patients (red line).

**Supplemental Figure 3**.

Plot of relative risks (RR) of in-hospital outcomes comparing epoch 4 patients to epoch 1-3 patients. The filled symbols represent the RR estimate; the lower and upper end of the vertical lines represent the lower and upper limits of the 95% confidence interval. Symbol interpretation is as follows: rectangle (unadjusted estimate); diamond (propensity score adjustment as a continuous variable); triangle (propensity score with inverse probability weighting (IPW)); circle (1:1 matched propensity score). Composite: in-hospital mechanical ventilation/mortality, LOS3: length of stay ≤3 days; LOS5: length of stay ≤5 days; LOS7: length of stay ≤7 days.

## Notes

**Conflict of Interest Disclosure:** None of the authors received any payments or influence from a third-party source for the work presented, and none report any potential conflicts of interest.

### Competing Interest Statement

The authors have declared no competing interest.

### Clinical Trial

Project was not registered as it was a retrospective Quality Improvement project approved by the UPMC Quality Improvement Review Committee

### Funding Statement

None.

### Author Declarations

The study received formal ethics approval by the UPMC Ethics and Quality Improvement Review Committee (Project ID 2882), the ethics/oversight body for ensuring patient confidentiality and consent (including waiver of consent) for analysis and dissemination of deidentified data within the UPMC system.

